# Magnitude, Associated Factors and Immediate Outcomes of Non-Reassuring Fetal Heart Rate Status Among Laboring Mothers at South Gondar Zone Public Hospitals, North, West Ethiopia, 2022; Cross Sectional Study

**DOI:** 10.1101/2022.10.02.22280615

**Authors:** Ewunetu Belete, Yibelu Bazezew, Melaku Desta, Dawit Misganaw, Mitiku Tefera

**Affiliations:** Ebinat Primary Hospital, Amhara Regional Health Bureau, Ethiopia; Department of Midwifery, College of Health Sciences, Debre Markos University, Debre Markos, Ethiopia; Department of Midwifery, College of Health Sciences, Assosa University, Assosa, Ethiopia; Department of Midwifery, Debre Birhan Health Science College, Amhara Regional Health Bureau, Ethiopia

**Keywords:** Debre Tabor, Ethiopia, laboring mothers, Non-reassuring fetal heart rate status

## Abstract

**Background:** Non-reassuring fetal heart rate status (NRFHRS) is an abnormal fetal heart rate monitoring which necessitates immediate intervention. It is one of the common reducible causes of perinatal morbidity and mortality in developing countries. Despite there is limited data on the magnitude, associated factors and its outcomes in Ethiopia.

**Objective:** To assess the magnitude, associated factors and immediate outcomes of non-reassuring fetal heart rate status among laboring mothers at South Gondar zone public hospitals, northwest Ethiopia 2022.

**Methods:** An institutional-based cross-sectional study was conducted from June 1-30, 2022. A total of 586 laboring mothers were included. The participants were selected through systematic sampling method. Bivariable and multivariable logistic regression analysis were carried out. OR with 95% CI was used and statistically significant variables were declared if p < 0.05 in multivariable analysis.

**Result:** The magnitude of NRFHRS was 21.16% (95%, CI: 17.9-24.7) with a response rate of 97.34%. Primigravida [AOR= 1.86, 95% CI: 1.03-3.37], anemia [AOR= 4.59, 95% CI: 1.87-11.30], referred [AOR= 1.95, 95% CI: 1.07-3.55], induction of labor [AOR= 3.78, 95% CI: 1.20-11.9], meconium-stained amniotic fluid [AOR= 14.13, 95% CI: 7.53-26.50], prolonged rupture of membrane [AOR= 11.70), 95% CI: 5.40-25.34] and low birth weight [AOR=5.08, 95% CI: 2.20-11.74] were significantly associated with NRFHRS. 4.8% of fetus was still birth.

**Conclusion:** In this study the magnitude of NRFHRS was high compared to studies in Africa. Being primigravida, anemic, referred, induction of labor, meconium-stained amniotic fluid, prolonged rupture of membrane and low birth weight were significantly associated with NRFHRS. Assigning adequate number of midwifes for good labor follow-up, properly counsel on nutrition and iron and folic acid and give due attention on labor follow-up would minimize NRFHRS.

## INTRODUCTION

Non-reassuring fetal heart rate status (NRFHRS) is defined as abnormal fetal heart rate monitoring, including irregular fetal heart rate (FHR) tone (fetal tachycardia or bradycardia), abnormal variability (no, minimal and/or marked variability), repeated fetal heart rate deceleration (late deceleration, variable deceleration and prolonged deceleration) (1, 2). NRFHRS is consider when a FHR is below 110bpm or FHR above 160bpm (1, 3). But according to Ethiopian Federal Ministry of Health (FMoH), NRFHS is diagnosed when a FHR is below 100bpm or above 180 bpm which lasts more than 10 minutes during labor and delivery for a normal (normal weight and term gestational age (GA) (4).

FHRS changes swiftly during labor, going from normal to abnormal and vice versa (3). Within physiological limits, a normal fetus can deal with the trauma of childbirth. But NRFHRS can emerge suddenly in a weakened fetus and/or an abnormal type of labor when the baby cannot adjust to changes in oxygen supply throughout the delivery process and also may be the cause by irregular or unsettling fetal heart rate during labor (5). FHR monitoring during labor and delivery is important to assess fetal wellbeing and to predict the outcome. The two most frequent methods of FHR monitoring are intermittent auscultation (IA) using (fetoscope and Pinard) and hand-held Doppler ultrasound, which are called electronical fetal monitoring (EFM) (6).

According to National Institute of Child Health and Human Development (NICHD), FHRS are categorized in three tier system for the sake of management, Category I includes baseline heart rate 110–160 bpm, moderate variability, and no late or variable decelerations, but accelerations and early decelerations may be present or absent. Category II, indeterminate FHRSs that are not Category I or III or no induced accelerations after fetal stimulation and Category III includes sinusoidal FHRP and absent baseline FHR variability followed by recurrent late decelerations, bradycardia, or recurrent variable decelerations (7, 8). Those categories are used only for facilities they have CTG, Federation of Gynecology and Obstetrics (FIGO) further recommends intermittent auscultation users for evaluating and categorizing FHRS in corporate to the case definition when CTG is unavailable (2).

Intrapartum FHR monitoring is a fundamental component of assessing fetal well-being during labor. Even though there were false positive rate to recognize NRFHRS, the affected fetus can lead to many health risks and injuries (9).

The magnitude of NRFHRS in the world lies between 8.9% to 30.7%, which causes approximately half of all stillbirths (10, 11) and there were an estimated 1.2 million stillbirths and 2.1million early neonatal deaths, among those 98% of these occurring in poor and middle-income countries, 77% in sub-Saharan Africa and south Asia (12-14).

NRFHS is a high-risk obstetric syndrome that has been related to an increased risk of perinatal morbidity, mortality (15) and related neonatal intensive care unit (NICU) admission (16). Furthermore the most common indication for instrumental deliveries, 36% (17) and 47.1%-58% for Vacuum and c/s respectively globally (18, 19).

Many medical and obstetrics factors like antepartum haemorrhage (APH), amniotic fluid disorders, induction/augmentation of labor, intrauterine growth restriction (IUGR), use of anesthesia, presence of MSAF, abnormal fetal presentation and intraoperative findings like (uterine rupture, cord entanglements or placental abruption were the risk factors identified to have association with NRFHS (10, 20, 21).

NRFHRS can leads to many health problems for newborn such as Hypoxia/anoxia, bad mood, frequent crying for a long duration, and intense crying (22), partial or total brain damage, cerebral palsy, paralysis, nerve damage(23), hearing and vision impairment, seizures, necrotizing enterocolitis, epilepsy, mental retardation, abnormal physical growth and perinatal death or stillbirth (15) and also increased risk of death of pediatrics through infection due to prolonged hospital stay (24).

NRFHRS tracing was described as requiring necessitates immediate intervention like caesarean section or instrumental vaginal delivery (11, 21), intrauterine resuscitation, including oxytocin deprivation and amnioinfusion (20) to prevent or reduce bad neonatal outcome. But determining the most effective strategy in the case of abnormal FHR tracings is so difficult. And also, there is still limited information on this important issue of abnormal FHR pattern (10).

In Ethiopia, to reduce the complication different strategies and policies were implemented to increase access of quality maternal, newborn and child health care like (Basic emergency obstetric and newborn care (BEmONC), comprehensive emergency obstetric and newborn care (CEmONC) and catchment based reproductive, maternal, newborn and child health (RMNCH) mentoring were implemented. Despite these newborn mortality remains a problem in Ethiopia, which is 30/1000 live birth reported. The Amhara region had the most highest (47/1000) deaths recorded (25). The majorities of the deaths were caused by abnormal fetal heart rate which can be minimized by strict intrapartum follow up (26). Therefore, this study needs to assess the magnitude, associated factors and immediate outcomes of non-reassuring fetal heart rate status among laboring mothers in south Gondar zone public hospitals, North West, Ethiopia.

## Methods and materials

### Study area

South Gondar is a zone in the Amhara region. Debre Tabor is the capital town of South Gondar, around 613 km north of Addis Ababa, 104 km northeast of Bahir Dar, 100 km southeast of Gondar and 50 kilometers east of Lake Tana.

South gondar is bordered on the south by east gojjam zone, on the southwest by west gojjam zone and bahir dar, on the west by lake tana, on the north by central gondar zone, on the northeast by wag Hemra zone, on the east by north wollo zone, and on the southeast by south wollo zone, with the abay river separating the two gojjam zones.

According to the zonal health depot 2019 report, there are 13 rural districts and 5 city administrations. Concerning to health service institutions there are one governmental referral hospital, nine primary hospitals (only 8 of which are fully functional), 73 health centers and 145 health posts.

### Study design and population

Institutional based cross-sectional study was conducted at South Gondar zone public hospitals from June1 - June 30/2022.

All laboring mothers who were giving birth in study area were source population and all laboring mothers who gave birth at south Gondar zone public hospitals during the study period were study populations.

A laboring mother diagnosed as intra-uterine fetal death (IUFD) on or before admission and a laboring mothers diagnose with gross congenital malformation (anencephaly, spinal bifida and hydrocephalus) were excluded in the study.

### Sample size determination

Sample size was determined by using a single population proportion formula by taking 95% confidence interval, 3% desire precision, proportion of NRFHRP 15.1% (at Finote Selam) (27).

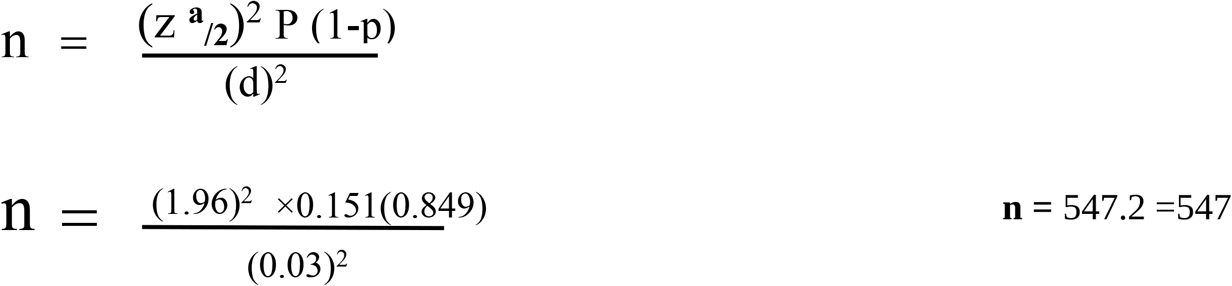

Then after by adding 10% non-respondent rate the final sample size was 602.

### Sampling procedure

There were 1 comprehensive specialized hospital and 8 primary public hospitals in south Gondar zone. By considering each hospital average monthly delivery report, total delivery in south Gondar zone hospitals were 1370. Then after, systematic random sampling was used, to select the study participants those laboring mothers who were came for birth during data collection until desired sample reached.

### Operational definitions

#### Non-reassuring fetal heart rate status (NRFHRS)

Considered when one or more than of the following (Tachycardia or bradycardia for more than 10 minutes), reduced FHR variability, decelerations and absence of accelerations) occur in the intrapartum period.

#### A baseline fetal heart rate status

normal between 110 to 160 bpm, Tachycardia >160 bpm, bradycardia <110 bpm (28).

#### Variability

Refers to the oscillations in the FHR signal, which are measured in 1-minute intervals as the average bandwidth amplitude of the signal includes normal variability (5-25 bpm), reduce variability (< 5 bpm) and increased variability (> 25 bpm).

#### Decelerations

FHR drops below baseline, with amplitude of more than 15 bpm and duration of more than 15 seconds. It can be (Variable decelerations that exhibit a rapid drop (onset to nadir in less than 30 seconds), Late decelerations delayed beginning and gradual return to baseline, as well as reduced variability and Prolonged decelerations that continue longer than 3 minutes (7).

#### Immediate birth out comes of NRFHRS

outcomes includes (APGAR score at 1^st^ minute and 5^th^ minutes, still birth, need for ventilation (asphyxia) and needs for neonatal NICU admission) (29), within five minute after delivery.

Unfavorable NRFHRS outcome: considered one or more the following (5-minute Apgar score less than 7, need for resuscitation in the delivery room, need for NICU, stillbirth (30)

#### Anemia

Defined as according to (WHO, 2011) (hemoglobin levels of ≥11 mg/dl was considered as normal, <11 mg/dl considered as anemia (31).

#### MUAC

according FOMH Ethiopia, 2022 Antenatal Care guideline ≥23 cm considered as normal <23 cm considered as malnourished (32).

### Data collection tools and procedures

A Structured and pretested questionnaire adapted from related literatures was used (27, 33). Data were collected through chart review and face-to-face interview. The questionnaire was prepared in English language and translated to the Amharic language and then translated back to English. Its consistency was checked by translating it back to English by different individuals. Nine diploma graduates in midwifes for data collectors and 3 BSc midwifes for supervisors were assigned. Data collection was conducted for one month from June 1-30/2022. The data collection process was supervised by the supervisors and principal investigator.

### Data processing and analysis

After data collected, the data was checked manually for completeness and consistency, and then entered in to Epi-data version 4.6.0.2 Software data entry template. Then data was exported to SPSS version 25 Software for analysis. Descriptive statistics was conducted and presented using frequencies, percentages, mean and standard deviation. Binary logistic regression analysis was employed to determine association between the dependent variables and the independent variable. Those variables with p-value of less than 0.25 on bi-variable were transferred to multivariable logistics regression for further regression analysis. Odd ratio with a 95% CI was computed to determine the strength and presence of association and variables with p-values of < 0.05 on multivariable were considered as statistically significant factors for NRFHRS. The backward stepwise regression model was used to enter variables in to model and the model fitness was checked using Hosmer-Lemeshow goodness of fit test and it was found (0.44) fit, and multicollinearity test was checked by Variance inflation factor (VIF), which was (1.02-3.13).

### Data Quality Control

To assure the quality of data, before actual data collection one day training was given on the objective of the study, procedures and techniques of data collection for the data collectors and supervisors by the principal investigator, Pretest was done on 5% of the total study eligible subjects at settings other than among the actual settings. Then the questionnaire was modified based on finding and the reliability of questionnaires. During data collection, the completeness, accuracy and consistency of collected data was checked daily during the data collection by supervisors and principal investigator.

### Ethical considerations

Ethical clearance was obtained from Ethical Review Committee (ERC) of Debre Markos University College of Health Sciences. Permission was also obtained and support letter to conduct the study was written from each hospital CCOs for Gyn/OBs department head of each hospital. And verbal informed consent (ascent age less than 18) was obtain from each laboring mother’s/attendant’s to obtained data and inform them for the purpose of the study, procedures, and their right to refuse or decline participation in the study at any time. Confidentiality also assured by telling all data were obtained from the chart and mothers were be kept confidential by using codes instead of any personal identities and is only for the purpose of the study.

## RESULTS

### Sociodemographic characteristics

A total of 586 laboring mothers included in this study with the response rate of 97.34%. More than one-third 217(37%) of the participant ages were found in the age category of 25-29 years, and the mean age was 26.8 years with SD of ± 5.41years. Most of participants were rural resident 325(55.5%). Around one third 206(35.2%) participants were farmers and 160(28%) of their husbands didn’t attend formal education **(Table 1)**.

**Table 1:** Socio-demographic characteristics of laboring mothers in South Gondar zone public hospitals, North West, Ethiopia 2022. (n=586)

| **Variables** | **Category** | **Frequency (%)** |
| --- | --- | --- |
| **Age** | 15-19 | 42(7.2) |
|  | 20-24 | 150(25.6) |
|  | 25-29 | 217(37.0) |
|  | 30-35 | 111(20.0) |
|  | >=35 | 66(11.3) |
| **Residence** | Rural | 325(55.5) |
|  | Urban | 261(44.5) |
| **Marital status** | Single | 9(1.5) |
|  | Married | 572(97.6) |
|  | Divorced | 5(0.9) |
| **Religion** | Orthodox | 551(94.0) |
|  | Muslim | 34(5.8) |
|  | Protestant | 1(0.2) |
| **Occupation** | House wife | 204(34.8) |
|  | Farmer | 206(35.2) |
|  | Gov't employ | 96(16.4) |
|  | Merchant | 63(10.8) |
|  | Dially laborer | 17(2.9) |
| **Women educational status** | Unable to read & write | 184(31.4) |
|  | Able to read and write | 65(11.1) |
|  | Primary education | 132(22.5) |
|  | Secondary education | 79(13.5) |
|  | Diploma and above | 126(21.5) |
| **Husbands' educational status (n=572)** | Unable to read & write | 160(28) |
|  | Able to read and write | 86(15.0) |
|  | Primary education | 88(15.4) |
|  | Secondary education | 80(14.0) |
|  | Diploma and above | 158(27.6) |
| **Women's MUAC** | <23cm | 180(30.7) |
|  | >=23cm | 406(69.3) |
| **MUAC=mid upper arm circumference** |  |  |

### Antepartum characteristics

Fifty-seven (14.8%) of the participants had experienced past obstetric problem and 114 (19.5%) had experienced history of past medical illness consecutively. Most, (93.9%) women had at least one ANC follow-up, but only 50% had >4 ANC follow-up. Ninety-six (16.4%) of women had recent pregnancy complication. Of thus, premature rupture of membrane (PROM) was the most prevalent current pregnancy complication (49.5%) **(Table 2)**.

**Table 2:**
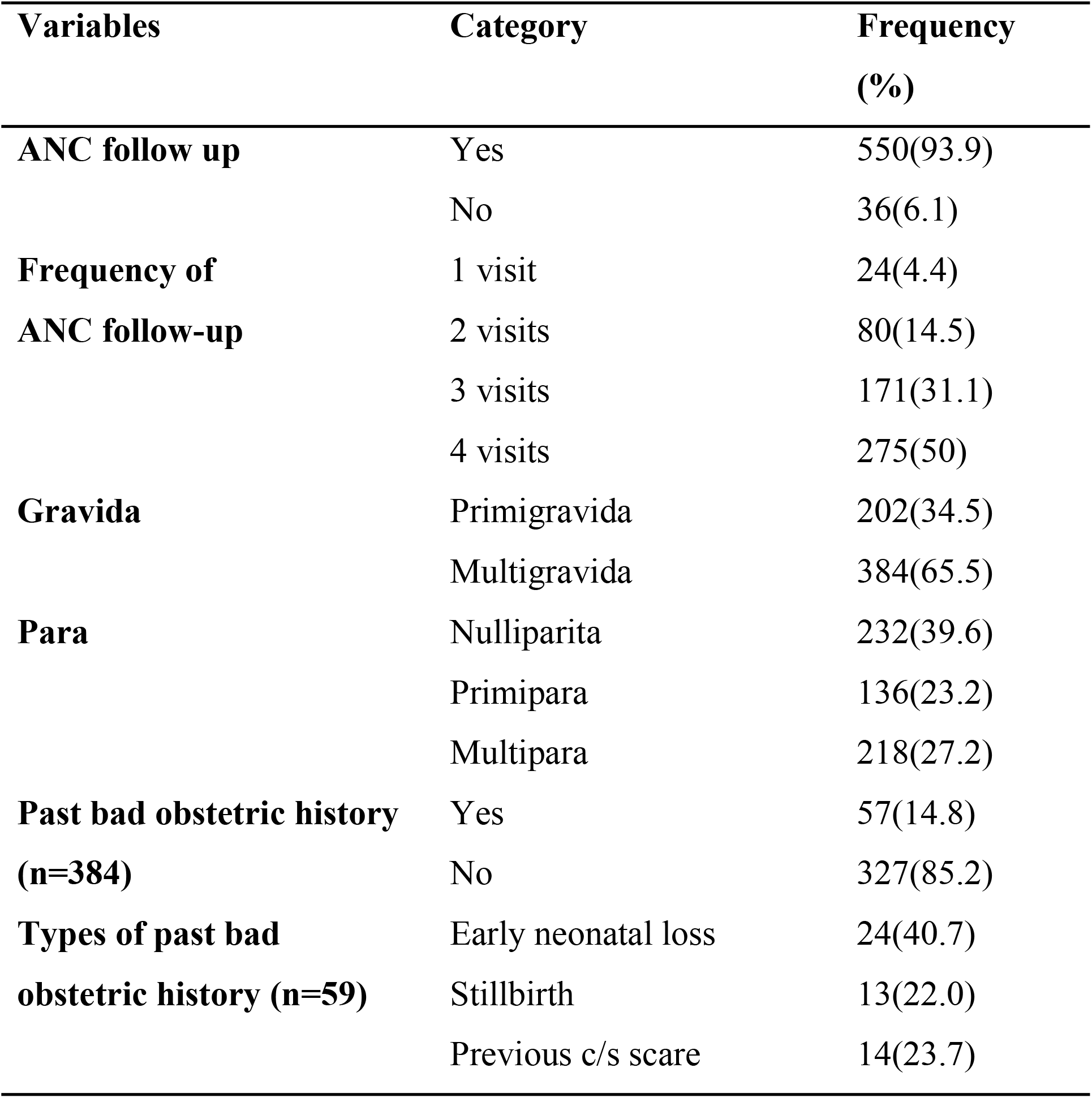
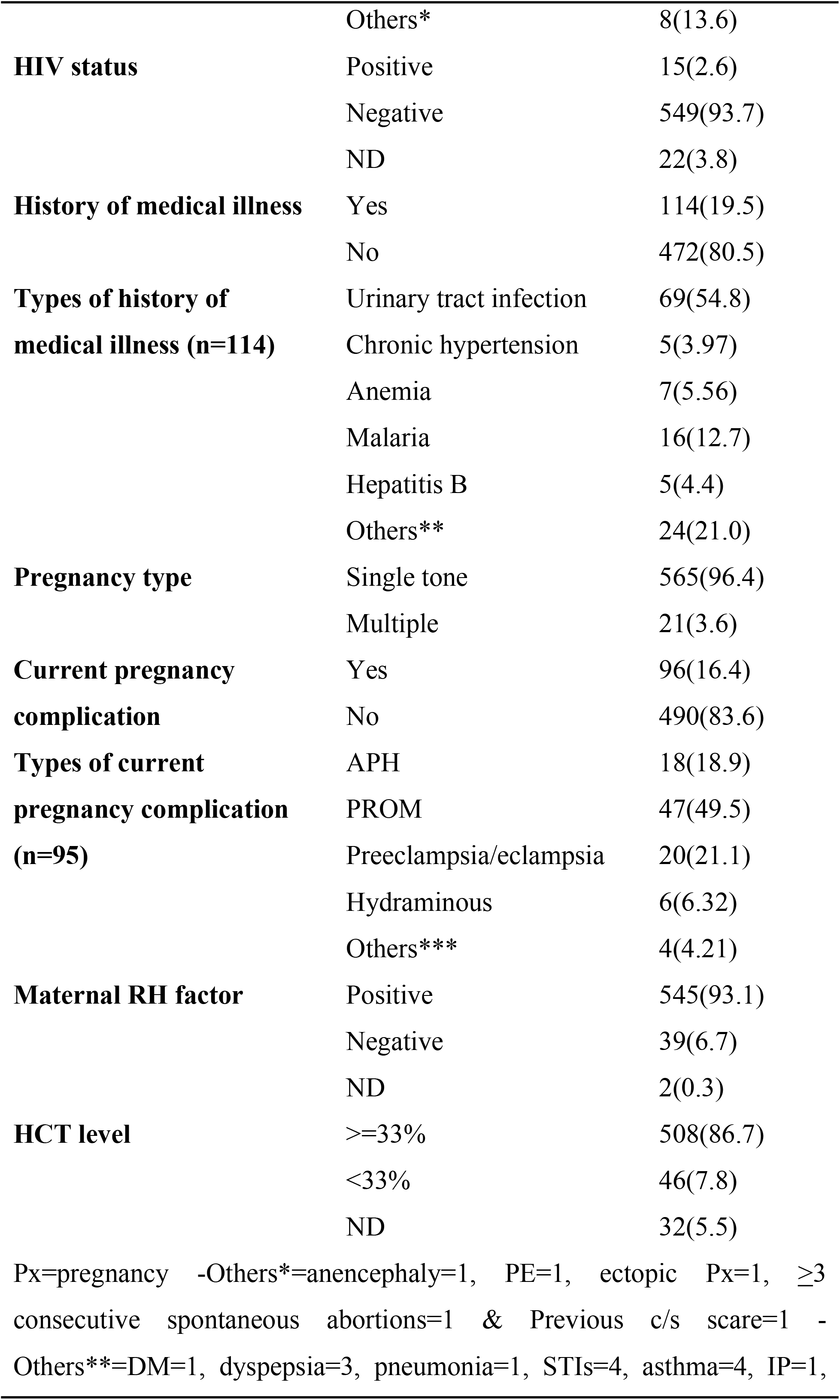
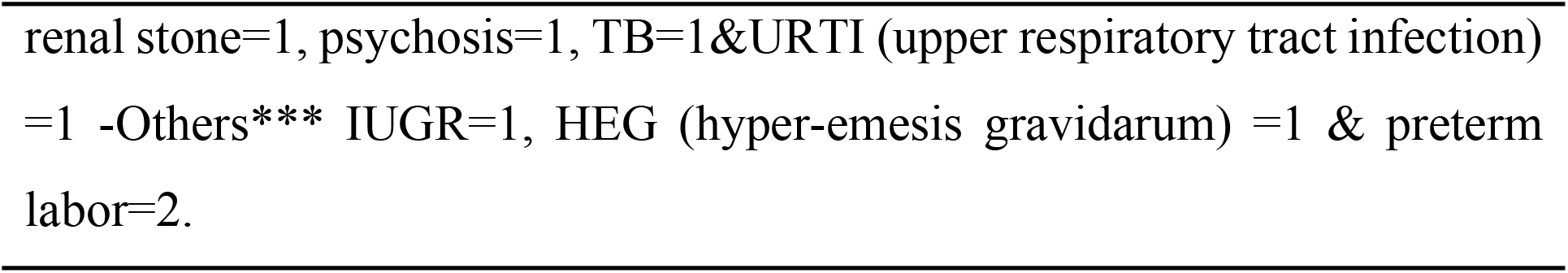
Antepartum characteristics of laboring mothers in south Gondar zone public hospitals North West, Ethiopia 2022 (n=586).

### Intra-partum characteristics

Four hundred twenty (71.7%) women know their last menstrual period and more than two third, 357(85%) of laboring mothers were 37-42 weeks of gestation. Two hundred one (34.3%) were referred from other health facility. More than half 303(51.7%) labors were latent first stage on admission and 127(21.7%) labor lasts greater than 18 hours. Again, 106(18.1%) laboring women had meconium-stained amniotic fluid. Most of neonates 505 (86.2%) had normal weight at birth with mean weight of 3056.16 gram ± 482.007, **(Table 3)**.

**Table 3:**
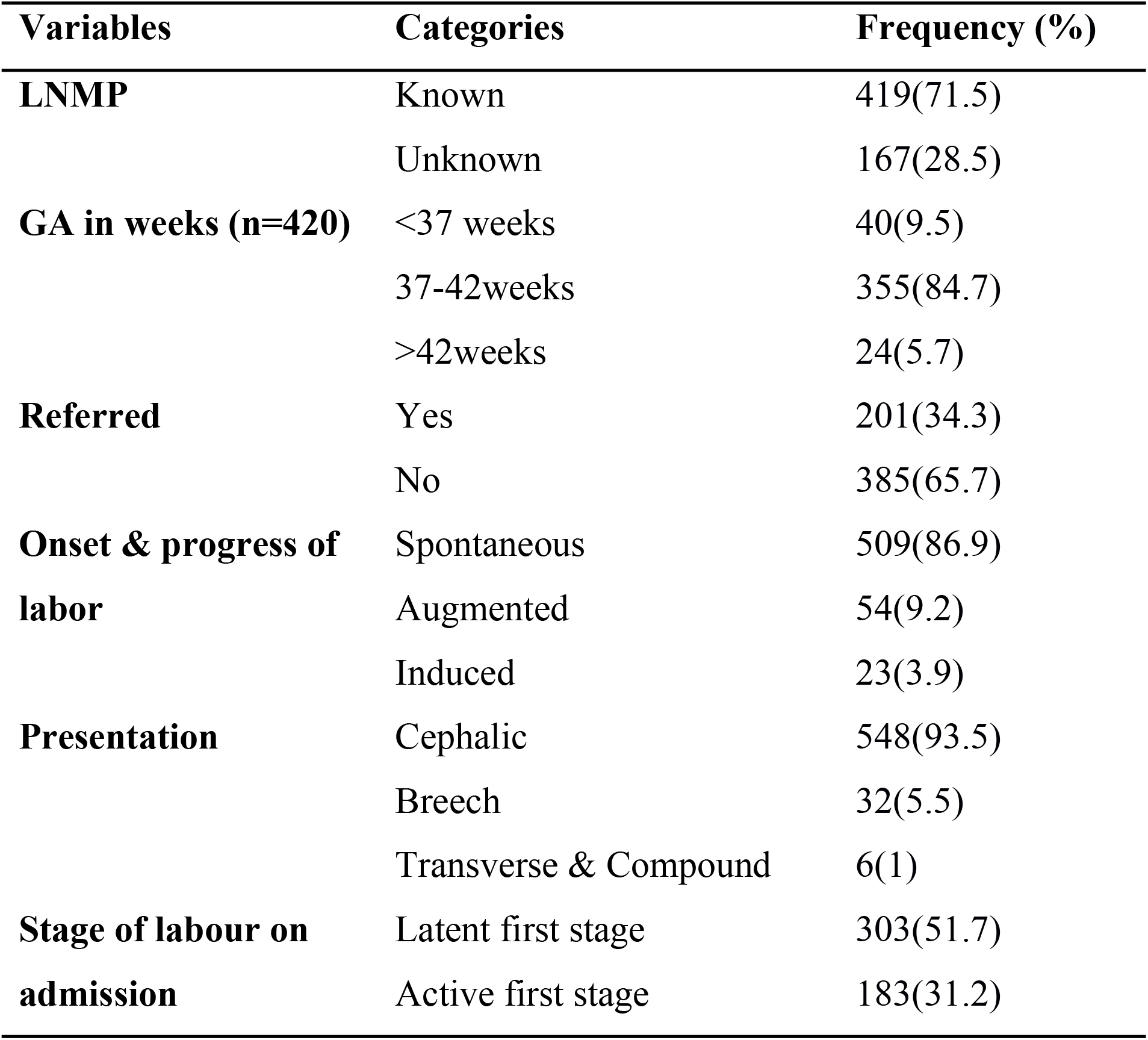
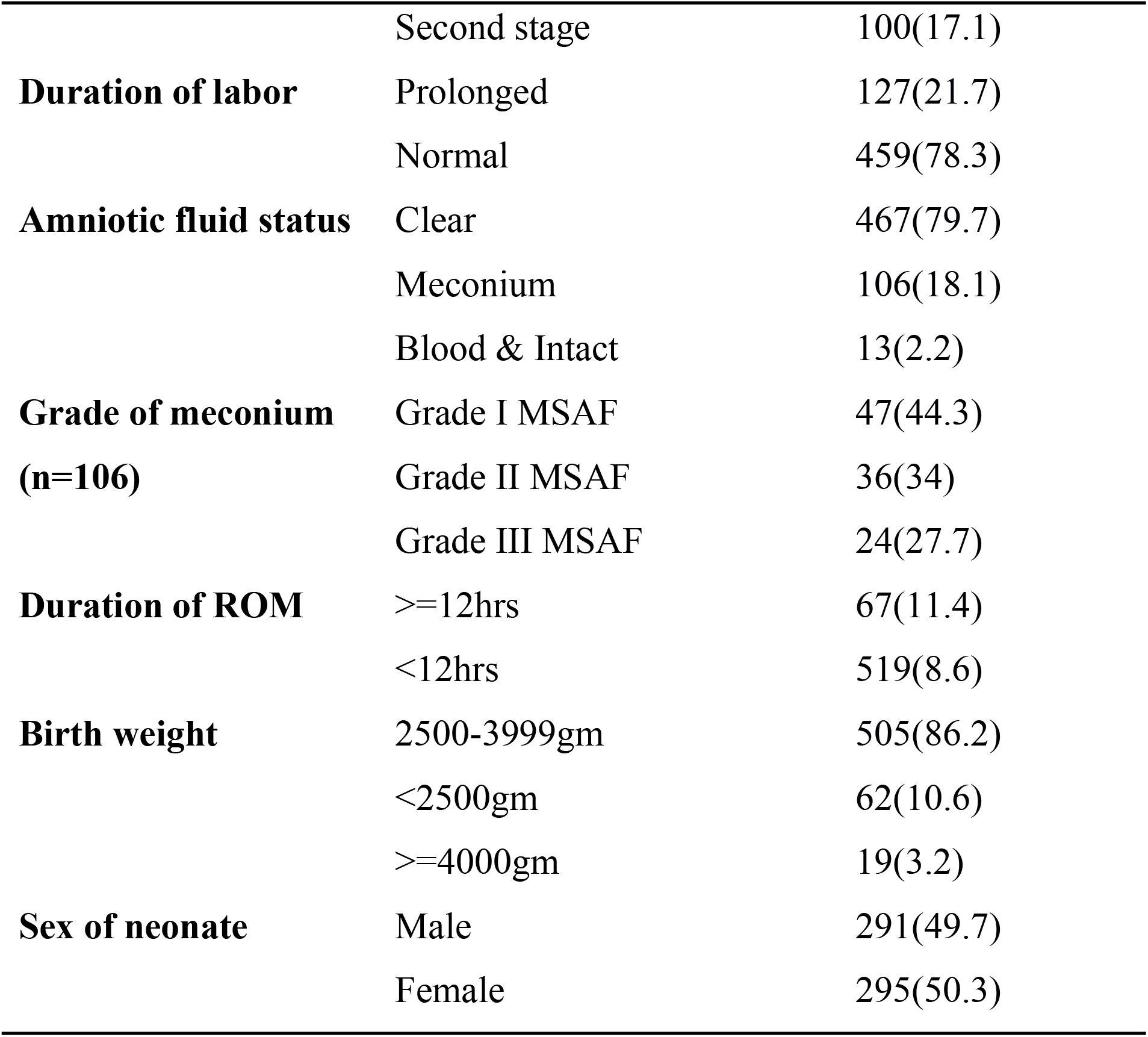
Intra-partum characteristics of laboring mothers in south Gondar zone public hospitals north west, Ethiopia 2022 (n=586).

### Magnitude of non-reassurance fetal heart rate status

More than one fifth (21.16%) with 95% CI: (17.9-24.7) labors were developed NRFHRS. Around half 63(50.8%) occur at second stage of labour and one third 45(34.1) caused by Nuchal (entanglement or knot) cord **(Table 4)**. More than one third 46 (37.1%) of NRFHRS fetus were delivered by c/s **(Fig 1)**.

**Table 4:**
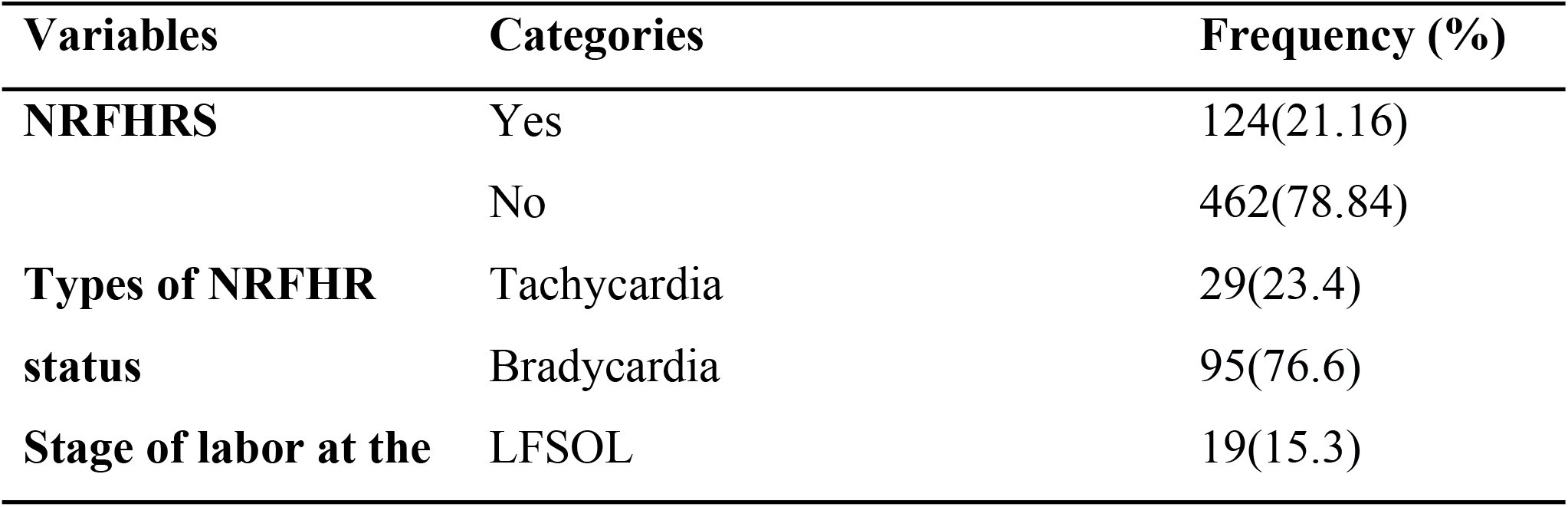
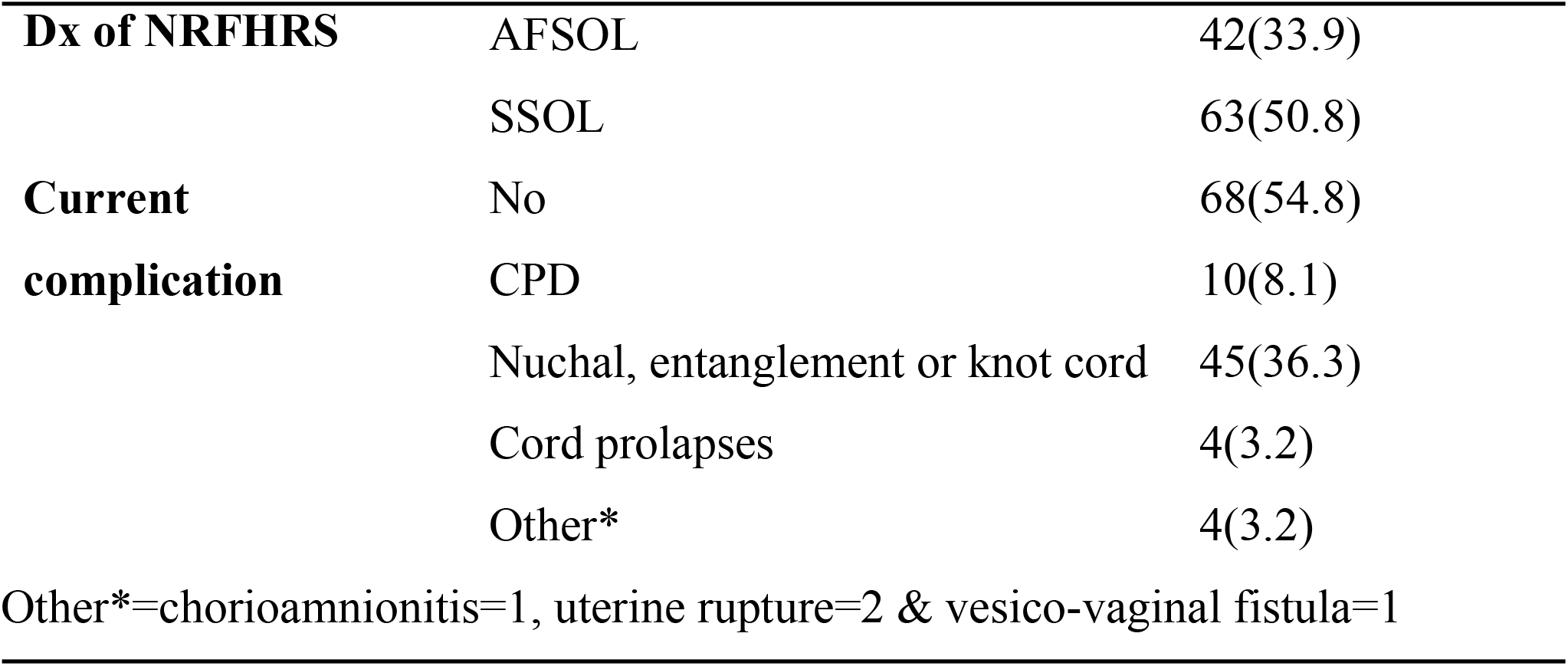
Magnitude and explanatory variables of NRFHRS among laboring mothers in south Gondar zone public Hospitals North West, Ethiopia 2022. (n=124).

**Table 5:**
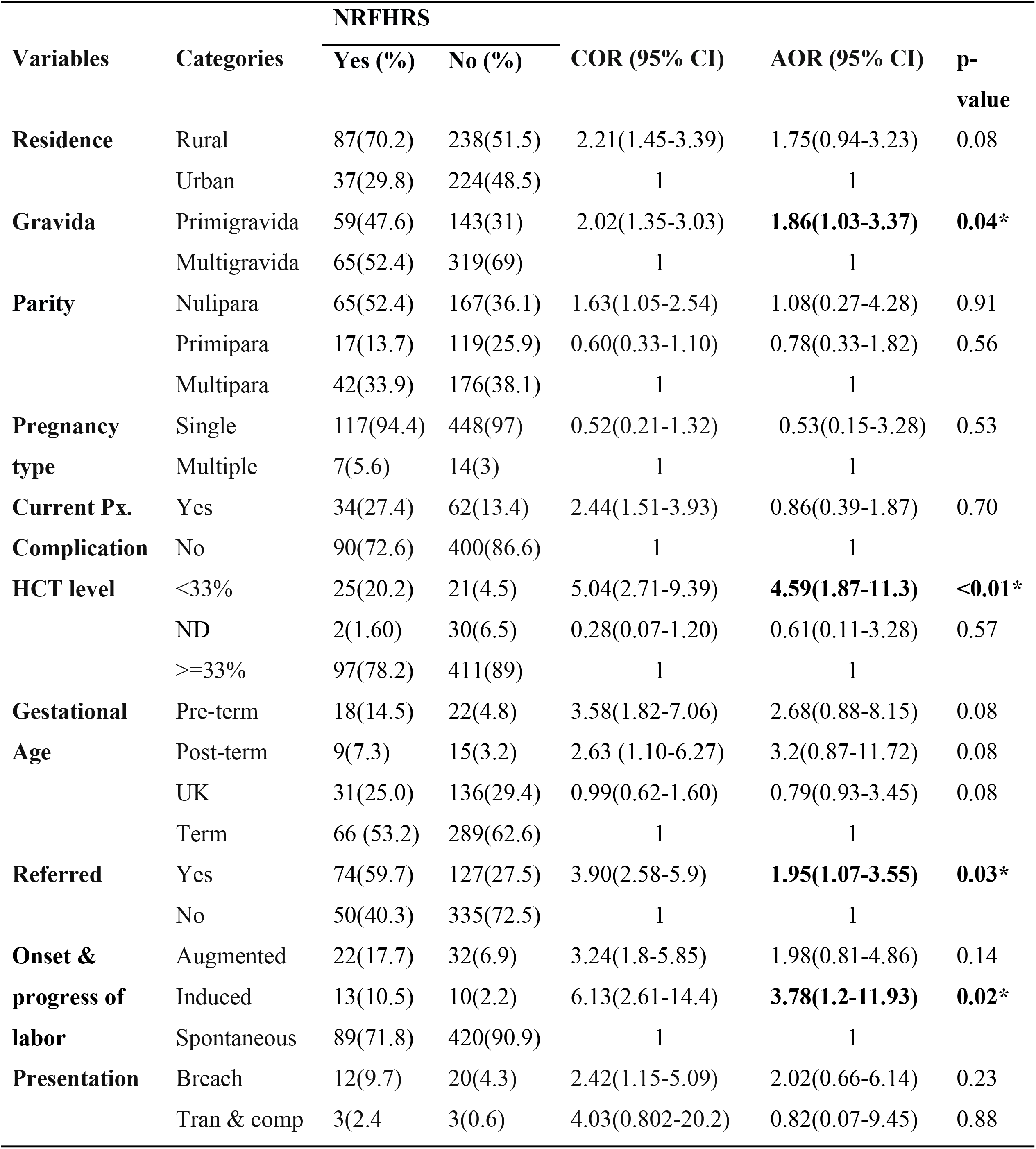
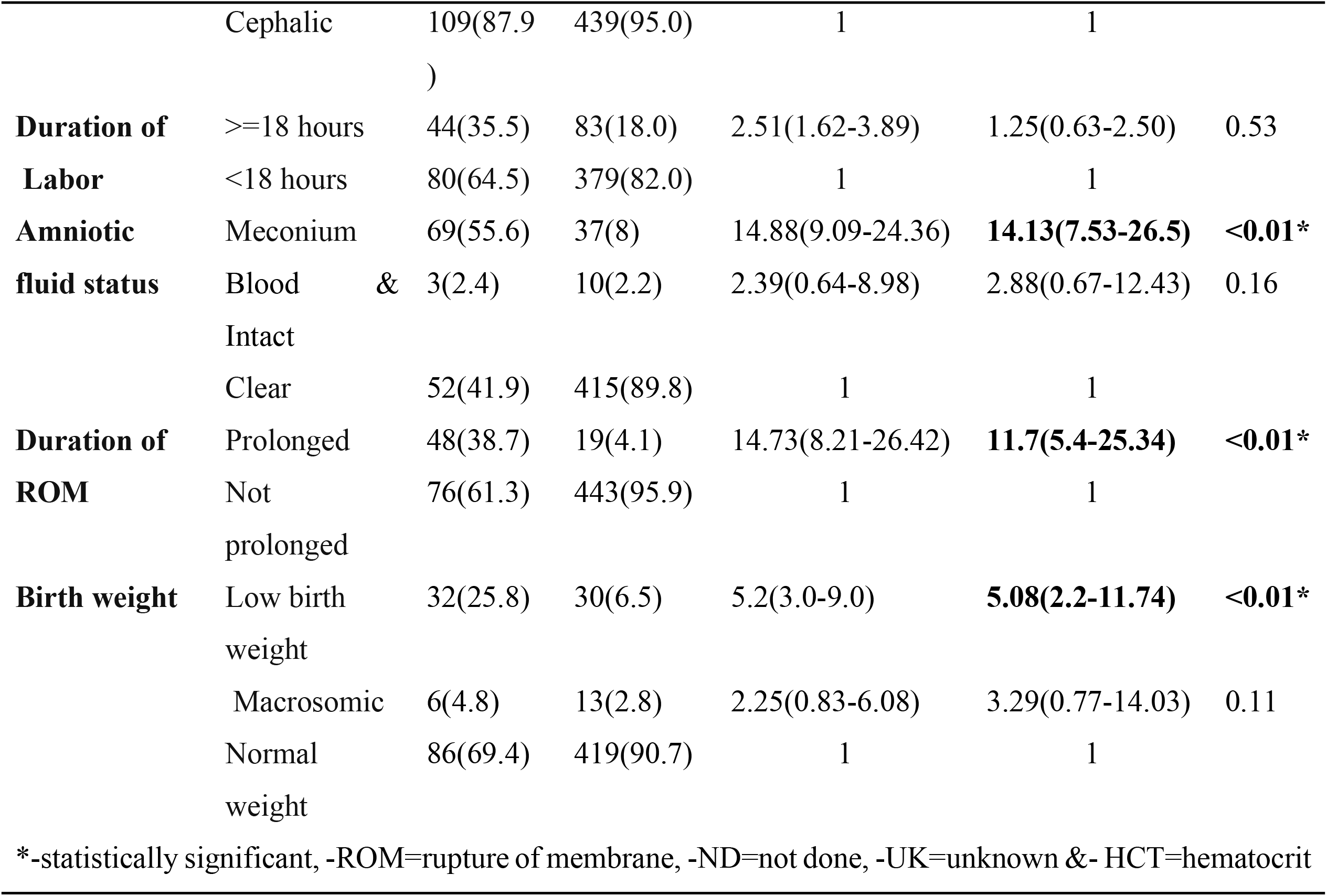
Bivariate and multivariable logistic regression analysis, factors associated with NRFHRS at south Gondar zone public hospitals, North West, Ethiopia 2022.

**Table 6:** Immediate outcomes of NRFHRS in south Gondar zone public hospitals north west, Ethiopia, 2022. (n=124).

| Variables | Categories | Frequency (%) |
| --- | --- | --- |
| **First minute APGAR score** | 0-3 | 9(7.3) |
|  | 4-6 | 52(41.9) |
| | $\geq 7$ | 63(50.8) |
| **Fifth minute APGAR score**<br>**(n=119)** | 0-3 | 2(1.7) |
|  | 4-6 | 19(16.0) |
| | $\geq 7$ | 98(82.4) |
| **Ventilation needed (n=118)** | Yes | 44(37.3) |
|  | No | 74(62.7) |
| **NICU admission needed**<br>**(n=118)** | Yes | 36(30.5) |
|  | No | 82(69.5) |
| **Still birth** | Yes | 6(4.8) |
|  | No | 118(95.2) |
| **Immediate NRFHRS outcome** | Favorable | 66(46.8) |
|  | Unfavorable | 58(53.2) |

**Fig 1:**
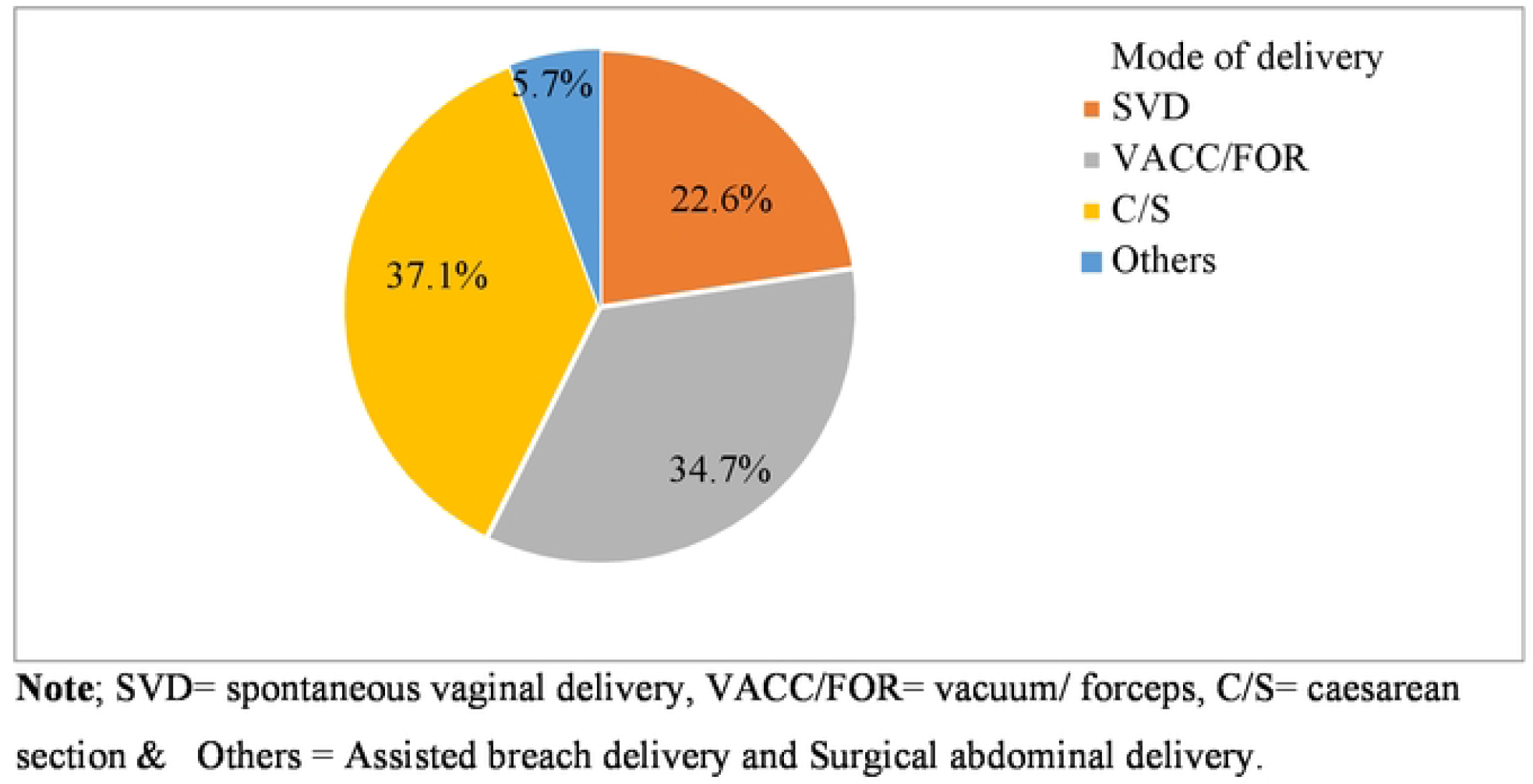
Mode of delivery of neonates after diagnosis of NRFHR in south Gondar zone public hospitals north west, Ethiopia 2022.

### Factors associated with NRFHRS

In this study, the association between socio-demographic characteristics, antepartum characteristics and intrapartum characteristics to NRFHRS was assessed. From all 14 variables which showed an association at the bivariable analysis were (residence, gravidity, parity, pregnancy type, current pregnancy complication, hematocrit level, gestational age, referred, onset of labor, presentation, duration of labor, duration of rupture of membrane, amniotic fluid status and birth weight). These variables (P-value <0.25) at Bivariable were transferred to and the multivariable analysis.

In multivariable analysis 7 variables (being primigravida, anemic, referred, induction of labor, meconium-stained amniotic fluid, prolonged rupture of membrane and low birth weight were showed significant associated with NRFHRS in the final model with 95% CI at p-values 0.05.

The odds of having NRFHRS among laboring women who were primigravida was increased by 86% (AOR= 1.86, 95% CI: 1.03-3.37) than among multigravida women. Similarly, the occurrence of NRFHRS in women who had anemia was 4.59 (AOR= 4.59, 95% CI: 1.87-11.3) times more likely than women those who were not anemic.

The odds of having NRFHRS among laboring women those who came from nearby health facility by referral was increased by 95% (AOR= 1.95, 95% CI: 1.07-3.55) than women those who came directly from home. Likewise, the odds of the presence of NRFHRS in women with induced labor was 3.78 (AOR= 3.78, 95% CI: 1.19-11.93) times more likely than women who had spontaneous onset of labor.

The odds of having NRFHRS was 14 times more likely (AOR= 14.1, 95% CI: 7.53-26.5) among women with meconium-stained amniotic fluid than those women whose amniotic fluid was clear. Also, the odds of NRFHS among women who had prolonged rupture of membrane was 11.7 (AOR= 11.7: 95% CI: 5.39-25.3) times more likely than their counter parts. This study also showed that odds of NRFHRS was increased by 5.08 (AOR=5.08, 95% CI: 2.20-11.7) times in low-birth-weight babies than normal birth weight babies (Table 7).

### Outcomes of NRFHRS

About half (50.8%) & more than three fourth (82.4%) NRFHRS fetus were delivered with APGAR score of greater than or equal to seven at first and five minutes. More than one third (37.3%) and around one third (30.5%) were needs ventilation and NICU admission after delivery respectively. And also, 4.8% of non-reassure fetus still birth. Regarding the overall neonatal outcome, nearly half 58 (46.8%) of neonates born unfavorable outcome, (Table 7).

## DISCUSSIONS

In this study the magnitude of NRFHRS was 21.16% (95%, CI: 17.9-24.7). This finding is in line with the studies conducted at Addis Ababa 18.6% (33) and Israel 21.2% (20). This might be due to the similarity of nearly equal sample size and socio demographic characteristics and study design. For instance, the age of participant in this study was similar with that of in the A.A most (37%) of participants were age category from 25-29 years both studies, unknown in GA 27.6 % in A.A, in this study also 28.5% and almost equal number of participants were LFSL on admission in all studies.

This finding was higher than the studies found in Felege Hiwot Comprehensive specialized Hospital, 12.2% (34), Finote Selam General Hospital, 15.1% (27), Nigerian, 8.9% (11), and China, 11.5% (35). This may be due to more than half of participants were rural resident which might be referred from nearby facilities (34.3%), high proportion of nulliparity (39.6%), which further causes increased number of NRFHRS (10, 35). Again, it can be explained due to only 50% of women complete the recommended ANC visit which increased the risk of NRFHRS due to missed opportunity of early detection and management of abnormalities that might cause NRFHRS status.

Another possible explanation for discrepancy might be also due to deferent study population like include single tone pregnancy only in & FHCSH (34), only term pregnancy in China (35), excluded malpresentation and unknown gestational age in FHCSH (34) and used secondary data Finote Selam (27), time gap more than five years between the studies in Nigerian (11), difference in methodology in China (35) & Nigerian (11).

In the contrary the finding is lower than study done in Thailand 30.7% (10), The discrepancy might be due to difference of time gap of study conducted variation in characteristics and also in Thailand there may be use more sensitive material to diagnosis NRFHRS.

This study revealed that primigravida mothers were more likely to have NRFHRS than multigravida mothers. The association is supported by studies conducted in Finote Selam (27), India (36) and Thailand (10). This might be due to being primigravida mothers are new for course of pregnancy and labor leading those to stress and anxiety which causes psychological and physiologic disruption which might decreased blood flow to the fetus and cause uterine hypoxia (37). Primigravida can be a risk of several pregnancy related complications such as prolonged duration of labor, pregnancy induced hypertension and other abnormal pregnancy and labour conditions which disturbed both maternal and fetal conditions (37, 38). But, this finding contrasts to the study conducted in Israel (20). This might be due to in Israel’s study there might be good ANC follow up and they may be properly counseled on the feature of pregnancy, danger sign and birth preparedness and complication readiness plan.

Similarly, laboring mothers who were anemic was increased the odds of NRFHRS compared to those who were not anemic. This finding is supported other studies done in India)(39), FMoH(4) and (2). This might be due to anemia is complication of other obstetric problems like (pre-eclampsia, APH) and anemia also most common medical problem during pregnancy cause fetal distress, moreover anemia by itself its own complication (decrease oxygen delivery to the fetus, IUGR and oligohydramnios) (39).

The study found that laboring mothers who were referred from nearby health facilities were more likely to develop NRFHRS. This finding is also identified in study conducted at Finote Selam (27). This might be due to most referred laboring mothers were complicated cases and also, they come from more remote area which has poor access for transportation and delay to seek care, also visited health center might be far from the hospital, leads to prolonged labor.

Mothers who had undergone induction of labor were more likely to develop NRFHRS than mothers who had spontaneous onset of labor. This study supported by studies conducted in Israel (20, 24) and Finote Selam (27) showed that oxytocin augmentation increased the risk of NRFHR. Administration of oxytocin to stimulate and hasten the labor process increase frequency of uterine contraction and decrease uterine blood flow it causes decrease blood and oxygen flow to the fetus and cause hypoxemia which change the FHR pattern (37). Moreover most of the time induction of labor was undergone from post term pregnancy, which also diminished utero placental blood flow, prone to oligohydramnios and meconium stained liquor lead to fetal hypoxia and fetal distress secondary to aged placenta (3, 40).

In this study amniotic fluid status was another factor for NRFHRS. Those mother whose amniotic fluid was meconium stained were more likely to develop NRFHRS than that had clear amniotic fluid, which is similar with other studies done India (38) and Finote Selam (27). This might be due to non-reassuring fetal heart rate increased risk of meconium stained amniotic vice versa (34) and presence meconium has an effect on umbilical vessels to constrict, which may lead to decrease blood and nutrients flow to the fetus (41) & and also MSAF increase risk of intrauterine endometrial inflammation (42) increased magnitude of NRFHRS. This finding contrast to study done in Punjab India (39). This may be due to the difference in study population and methodology, similarly the study done in India was only included term pregnancy with cephalic presentation, but in this study, includes fetuses that were in presentation other than cephalic, preterm and post-term gestations were included.

Moreover, the duration of rupture of membrane was significant factor for NRFHRS. Thus, mother who had prolonged duration of ruptured membrane were more likely to develop NRFHRS than their counterparts. This finding is similar with a study done in Israel (20). This may be due to prolonged duration of rupture membrane cause infection (chorioamnionitis) and sever oligohydramnios which increase tension of the fetuses.

This study also found that fetuses which had low birth weights were more likely to the odds of NRFHRS compared to fetuses that had normal birth weight. This finding is supported by a study conducted in Israel (20). The reason might be due to low birth weight fetuses are most of the time small for gestational age, and also LBW resulted from multiple pregnancy, abnormal pregnancy (pregnancy induced hypertension, antepartum hemorrhage, PROM) and preterm gestation which increased risk of NRFHRS (35).

Furthermore, this study revealed that around half of newborns with NRFHR status (49.2%) were 1^st^ minute APGAR score of less than 7. This finding is higher than the study done in 11.32% (15). This difference was might be due to methodology and time gap to the study. But this finding of this study is lower than the study done in Nigeria 1^st^ minute APGAR was 70.9%. The discrepancy may be due to in this study are most of neonate delivered through vacuum, because (80% neonatal outcome was not favorable birth given by vacuum compared to neonatal outcome than those with forceps and c/s deliveries (43).

About one third (30.5%) of NRFHRS fetuses require NICU admission after delivery in this study. This result is similar with the study conducted in Bangladesh (28.3%) (15). The similarity may be due to methodology and might be similarity of demographic though not described.

In this study neonates born after developed NRFHRS were 37.3% of neonates were require ventilation immediately after delivery. This is higher than the finding of studies done at Rwanda (17.2%) (44) and Tanzania (6.1%)(12). The reason may be due to they might be used more sensitive material to diagnosis NRFHRS and take immediate action soon after the diagnosis in Rwanda and Tanzania.

In present study, 4.8% fetuses were still birth from after developed NRFHRS. This finding is higher than the studies done in Rwanda 1.1%(44), Nigeria 3.125% (11) and Tanzania 2.5% (12). The difference might be due to there were used CTG than in this area because CTG was highly sensitive (45) to diagnosis NRFHRS and also taken appropriate action as soon as immediately like C/S.

### Limitation of the study

We can’t do anything without limitation and challenge, there for this study has its own limitation. Even tracing of FHB was very challenging, laboring mothers followed till outcome occurred or delivered (once NRFHRS diagnosed considered as NRFHRS). And tracing of twin pregnancy was also anther challenge and there might be also subjectivity in the diagnosis of NRFHRS.

## CONCLUSION AND RECOMMENDATIONS

### Conclusion

The magnitude of NRFHRS is comparatively high when compared to studies done in Africa and associated with unfavorable birth outcome nearly half neonates. Being primigravida, anemic, referral from nearby health facility, induction of labor, meconium-stained amniotic fluid, prolonged rupture of membrane and low birth weight were significantly associated with NRFHRS. Nearly half of neonate effected unfavorable outcome.

### Recommendations

Zonal health depot and district health office: be ready ambulance service specially those who are remote area to reduce delay because being referred is risk factor. Strengthen the clinical mentoring this mentoring also alert professionals to identify problems as early as possible.

For health center and hospital midwifery: Consciously follow primigravida laboring mothers and give them adequate information about the nature of labor, psychological support and possible intrapartum complications they may face. Should be encouraged mothers to have the recommended number of ANC follow-up and properly take iron with folic acid supplementation to reduce anemia and give proper counseling on nutrition this also reduce risk of low birth weight. Strengthen communication with the referred and referring facility.

As MSAF showed strong association, giving due attention to its presence and anticipating NRFHRS may prevent further complications. During induction, of labor, it is better considered possible complications and monitor closely. Take immediate (corrective) action for prolonged rupture of membrane and meconium-stained amniotic fluid.

For researchers: I recommend, further study is needed by strong design and associated factors for outcomes, because in this study association was not done for the outcomes.

## Data Availability

The original data for this study are available from the corresponding author upon reasonable request.

## Acronyms and Abbreviations

APH: Ante-Partum Hemorrhage
CTG: Cardiotocogram
CS: Caesarean Section
EFM: Electrical Fetal Monitoring
FHR: Fetal Heart Rate
FMoH: Federal Ministry of health
HCT: Hematocrit
IUFD: Intra Uterine Fetal Death
IUGR: Intra Uterine Growth Retardation
MSAF: Meconium-Stained Amniotic Fluid
NICHD: National Institute of Child Health and Human Development
NICU: Neonatal Intensive Care Unit
NRFHR: Non-Reassuring Fetal Heart Rate
PROM: Premature/Prelabor Rupture of Membrane
ROM: Rupture of Membrane
RMNCH: Reproductive Maternal and Child Health
SPSS: Statistical Product and Service Solution

## Acknowledgments

First, we would like to acknowledge the Debre Markos University Department of Midwifery, Amhara Regional Health Bureau, hospital medical directors and labor & delivery ward coordinators, all the data collectors and study participants.

## Author Contributions

Conceptualization: EB, YB, MD, DM, MT. Formal analysis: EB. Funding acquisition: EB. Investigation: EB, YB, MD, DM. Methodology: EB, YB, MD, DM, MT., Supervision: EB, DM. Validation: EB, YB, MD, DM, MT. Visualization: EB, YB, MD, DM, MT. Writing-original draft: EB, DM. Writing-review& editing: EB, YB, MD, DM, MT.

